# Impact of Fludrocortisone on the Outcomes of Subarachnoid Hemorrhage Patients: A Retrospective Analysis

**DOI:** 10.1101/2023.09.28.23296246

**Authors:** Akshitkumar M. Mistry, Janki Naidugari, Michael J. Feldman, Jordan A. Magarik, Dale Ding, Isaac J. Abecassis, Matthew W. Semler, Todd W. Rice

## Abstract

**Background:** Whether the use of fludrocortisone affects outcomes of patients with aneurysmal subarachnoid hemorrhage (aSAH) and its usage rate in the United States remain unknown.

**Methods:** We conducted a retrospective analysis of 78 consecutive patients with a ruptured aSAH at a single academic center in the United States. The primary outcome was the score on the modified Rankin scale (mRS, range, 0 [no symptoms] to 6 [death]) at 90 days. We adjusted the primary outcome for age, hypertension, aSAH grade, and time from aSAH onset to aneurysm treatment. Secondary outcomes were brain and cardiopulmonary dysfunction events.

**Results:** Among 78 patients at a single center, the median age was 58 years [IQR, 49 to 64.5]; 64% were female, and 41 (53%) received fludrocortisone. The adjusted common odds ratio, aOR, of a proportional odds regression model of fludrocortisone use with mRS was 0.33 (95% CI, 0.14-0.80; P=0.02), with values <1.0 favoring fludrocortisone. Organ-specific dysfunction events were not statistically different: delayed cerebral ischemia (22% vs. 39%, P=0.16); cardiac dysfunction (0% vs. 11%; P=0.10); and pulmonary edema (15% vs. 8%; P=0.59).

**Conclusions:** The risk of disability or death at 90 days was lower with the use of fludrocortisone in aSAH patients.

## INTRODUCTION

Inpatient and overall mortality of patients with aneurysmal subarachnoid hemorrhage (aSAH) are 20% (1-3) and 40%, respectively (4). The high morbidity and mortality result from life-threatening neurological sequelae like re-hemorrhage, cerebral swelling, and vasospasm, which leads to ischemic strokes, as well as non-neurological complications like fatal volume loss from cerebral salt wasting (kidney-drive natriuresis), acute kidney injury (AKI), and cardiopulmonary injury (5-7).

Patients with aSAH can have fatal polyuria (>4 liters/day) from natriuresis (8, 9). Thus, they require incredibly large volumes of intravenous fluids to maintain their intravascular volume (8-10). Hence, fluid therapy is the most common therapy aSAH patients receive. In the presence of natriuresis-driven polyuria, fluid administration is among the most considerable challenges in the medical management of aSAH patients. Failure to do so can compromise multi-organ perfusion, increasing the risk of life-threatening ischemic strokes, cardiopulmonary dysfunction, and AKI (11).

Fludrocortisone is well-known to reduce natriuresis and hyponatremia, but its effects on outcomes in aSAH patients have not been studied. Using data from a prospective study and two phase 2 randomized controlled trials, we have shown that fludrocortisone decreases natriuresis, hyponatremia, and polyuria in aSAH patients, leading to lower fluid and sodium requirements and hypovolemic incidences (8). Importantly, a meta-analysis of the studies shows that it lowers the rate of symptomatic vasospasm (8). The 2023 American Heart Association aSAH guidelines recommend that “the use of mineralocorticoids is reasonable to treat natriuresis and hyponatremia” (12).

However, the impact of fludrocortisone on the clinical outcomes of aSAH patients has yet to be studied. Therefore, to fill this knowledge gap, we conducted a retrospective analysis of aSAH patients enrolled in the SMART randomized trial,(13) comparing outcomes of patients treated with fludrocortisone with those not treated with fludrocortisone. We hypothesized that, because of the established efficacy of fludrocortisone, its use would result in less death or disability, as measured by the modified Rankin Scale (14) (mRS), at 90 days.

## METHODS

### Study Design and Patient Selection

We conducted a retrospective cohort analysis of all aSAH patients enrolled in the Isotonic <u>S</u>olutions and <u>M</u>ajor <u>A</u>dverse <u>R</u>enal Events <u>T</u>rial (SMART)(13) after its results and data of aSAH patients were available. SMART was a pragmatic, unblinded, cluster-randomized, multiple-crossover clinical trial (ClinicalTrials.gov: NCT02444988, NCT02547779) conducted at an academic medical center in the United States between June 1, 2015, and April 30, 2017. Trial protocol did not control the use of fludrocortisone. This subanalysis was approved by the institutional review board at Vanderbilt University Medical Center (IRB # 141349).

### Data Collection

Data for aSAH patients were collected retrospectively blinded to fludrocortisone use. The following co-variables were collected: past medical history (hypertension, diabetes, ischemic heart disease, tobacco use, and prior aSAH); aSAH severity as measured on Hunt and Hess scale, the World Federation of Neurological Surgeons (WFNS) grading system, and the modified Fischer scale; location, largest linear measurement, and treatment procedure (endovascular embolization vs. clip obliteration) of the ruptured aneurysm; time from symptom onset to procedural intervention; new stroke (iatrogenic or spontaneous aneurysmal re-rupture); development of hydrocephalus requiring ventricular cerebrospinal fluid drainage; and infection of the central nervous system, meninges, or cerebrospinal fluid (microbiologically proven or clinical signs treated empirically with 4 or more consecutive days of antibiotics).

### Study Intervention: Fludrocortisone Administration

The general institutional practice was to maintain euvolemia in aSAH patients and treat electrolyte imbalances. There was no institutional protocol to maintain euvolemia and treat hyponatremia. The use of fludrocortisone (dosage regimen and length), enteral salt, and/or hypertonic saline was provider dependent. Fludrocortisone was frequently initiated in patients with vasospasm who were at risk of hypovolemia. In this cohort, fludrocortisone was started as early as the 3^rd^ day of ICU admission and maintained for the majority of ICU stay.

### Study Outcomes

Our study outcomes were not part of the SMART trial. Our primary outcome was the ordinal score on the mRS (14) (range, 0 [no symptoms] to 6 [death]) around 90 days. It was collected prospectively as part of routine clinical follow-up. Secondary outcomes included:

- the 21-day incidence of symptomatic vasospasm (defined as at least a 2-point decrease in Glasgow coma score or 2-point increase in the motor score of the National Institutes of Health Stroke Scale lasting for at least 8 hours after exclusion of attributable cerebral and non-cerebral conditions) and delayed cerebral ischemia (defined as a presence of a new, post-operative hyperintensity on diffusion-weighted magnetic resonance imaging, or a new, permanent focal neurological deficit, or a permanent decrease on Glasgow coma score of at least 2 points unattributable to other cerebral and non-cerebral conditions); (15, 16)
- total amount of fludrocortisone, enteral sodium chloride, and hypertonic saline (3% sodium chloride) received during the hospitalization;
- cardiac dysfunction defined as myocardial infarction, new onset of heart failure, or initiation of inotropic therapy;
- pulmonary edema evidenced by a chest radiograph or diuretic treatment with the goal of treating pulmonary edema.

### Statistical Analysis

Chi-square test for categorical data and the Wilcoxon test for continuous data were utilized for between-group comparisons without adjustment for covariates. The primary analysis compared 90-day mRS scores treated as an ordinal variable between patients treated with and without fludrocortisone using a proportional odds regression model, adjusting for age, history of hypertension, time from aSAH onset to aneurysm treatment, procedure received (clip or embolization), and the WFNS grade at baseline. This outcome was not assessed at the 90-day timepoint in 6 patients; thus for them, the last known mRS score was carried forward. To assess the robustness of the primary analysis, we performed three sensitivity analyses in which we repeated the primary analysis: 1) excluding patients with missing 90-day mRS scores; 2) substituting the mRS scores with values imputed using multivariate imputation by chained equations (mice v3.11 and mitools v2.4 R packages) in which one hundred different datasets were generated with imputed values and results were pooled; 3) any co-variable correlated with fludrocortisone was introduced into the model to adjust its effects; and 4) intervention tested in the SMART trial. The results of the regression models are presented with a point estimate odds ratio and associated 95% confidence interval (CI) range. Significance was set at a two-tailed α≤0.05. All analyses were performed using R version 4.0.2 software (R Foundation for Statistical Computing, Vienna, Austria).

## RESULTS

### Baseline Patient Characteristics

The median age of the 78 aSAH patients included in this study was 58 years (interquartile range, IQR, 49 to 64.5) and 50 patients (64%) were female. A total of 70 patients (90%) received an endovascular embolization of their ruptured aneurysm, 46 (59%) of which were in the anterior cerebral circulation. Forty-one patients (53%) were treated with fludrocortisone and 37 patients (47%) were not treated with fludrocortisone. Besides age, which was lower in the group treated with fludrocortisone, the two groups were similar at baseline (Table 1).

**Table 1.**
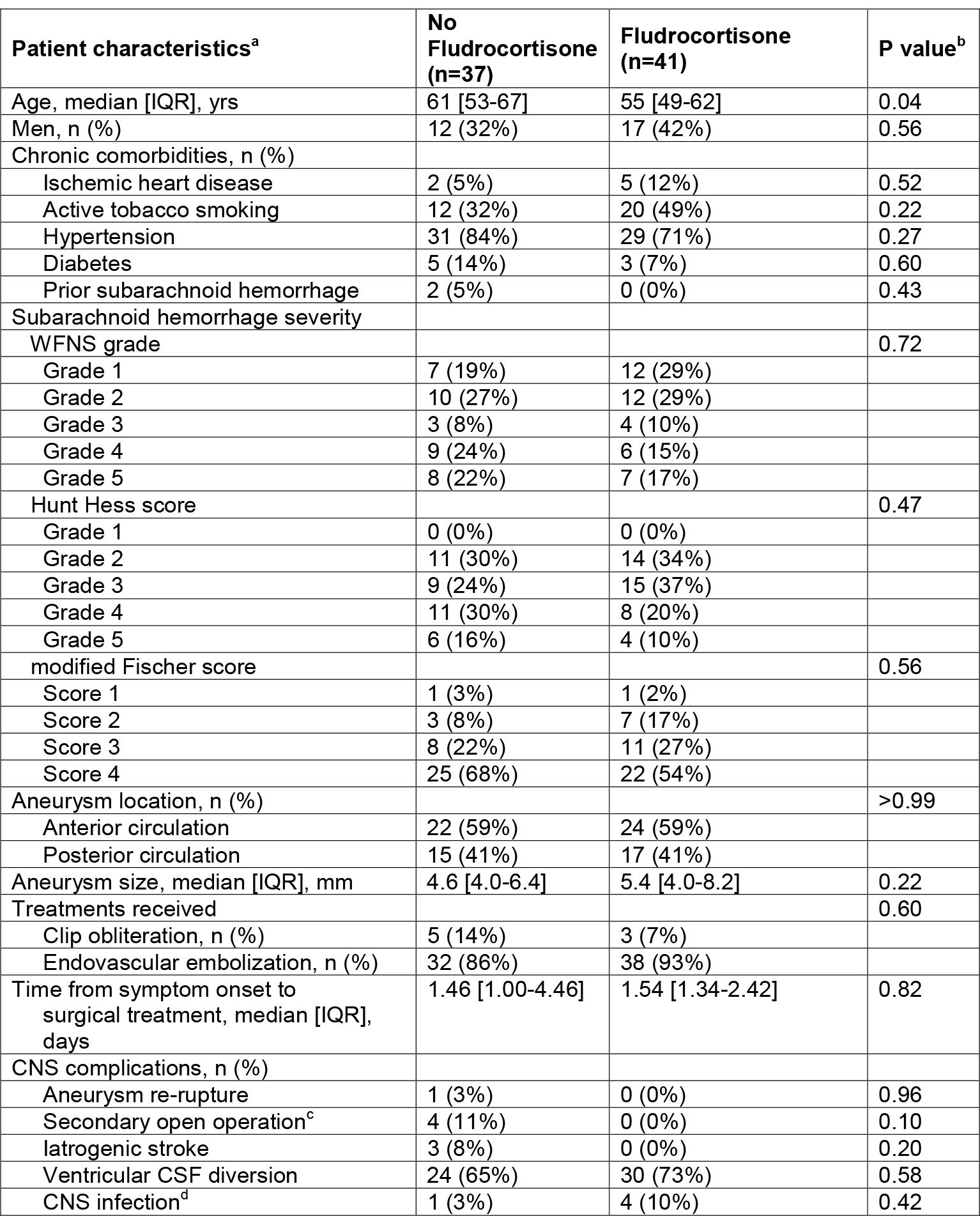
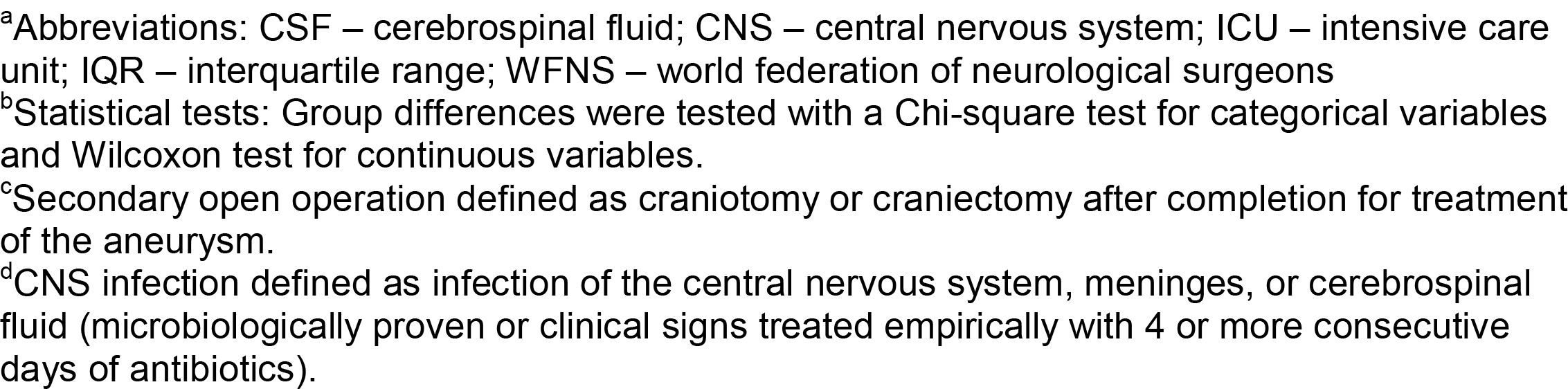
Patient characteristics at baseline.

| Patient characteristics<sup>a</sup> | No Fludrocortisone (n=37) | Fludrocortisone (n=41) | P value<sup>b</sup> |
| --- | --- | --- | --- |
| Age, median [IQR], yrs | 61 [53-67] | 55 [49-62] | 0.04 |
| Men, n (%) | 12 (32%) | 17 (42%) | 0.56 |
| Chronic comorbidities, n (%) |  |  |  |
| Ischemic heart disease | 2 (5%) | 5 (12%) | 0.52 |
| Active tobacco smoking | 12 (32%) | 20 (49%) | 0.22 |
| Hypertension | 31 (84%) | 29 (71%) | 0.27 |
| Diabetes | 5 (14%) | 3 (7%) | 0.60 |
| Prior subarachnoid hemorrhage | 2 (5%) | 0 (0%) | 0.43 |
| Subarachnoid hemorrhage severity |  |  |  |
| WFNS grade |  |  | 0.72 |
| Grade 1 | 7 (19%) | 12 (29%) |  |
| Grade 2 | 10 (27%) | 12 (29%) |  |
| Grade 3 | 3 (8%) | 4 (10%) |  |
| Grade 4 | 9 (24%) | 6 (15%) |  |
| Grade 5 | 8 (22%) | 7 (17%) |  |
| Hunt Hess score |  |  | 0.47 |
| Grade 1 | 0 (0%) | 0 (0%) |  |
| Grade 2 | 11 (30%) | 14 (34%) |  |
| Grade 3 | 9 (24%) | 15 (37%) |  |
| Grade 4 | 11 (30%) | 8 (20%) |  |
| Grade 5 | 6 (16%) | 4 (10%) |  |
| modified Fischer score |  |  | 0.56 |
| Score 1 | 1 (3%) | 1 (2%) |  |
| Score 2 | 3 (8%) | 7 (17%) |  |
| Score 3 | 8 (22%) | 11 (27%) |  |
| Score 4 | 25 (68%) | 22 (54%) |  |
| Aneurysm location, n (%) |  |  | >0.99 |
| Anterior circulation | 22 (59%) | 24 (59%) |  |
| Posterior circulation | 15 (41%) | 17 (41%) |  |
| Aneurysm size, median [IQR], mm | 4.6 [4.0-6.4] | 5.4 [4.0-8.2] | 0.22 |
| Treatments received |  |  | 0.60 |
| Clip obliteration, n (%) | 5 (14%) | 3 (7%) |  |
| Endovascular embolization, n (%) | 32 (86%) | 38 (93%) |  |
| Time from symptom onset to surgical treatment, median [IQR], days | 1.46 [1.00-4.46] | 1.54 [1.34-2.42] | 0.82 |
| CNS complications, n (%) |  |  |  |
| Aneurysm re-rupture | 1 (3%) | 0 (0%) | 0.96 |
| Secondary open operation<sup>c</sup> | 4 (11%) | 0 (0%) | 0.10 |
| Iatrogenic stroke | 3 (8%) | 0 (0%) | 0.20 |
| Ventricular CSF diversion | 24 (65%) | 30 (73%) | 0.58 |
| CNS infection<sup>d</sup> | 1 (3%) | 4 (10%) | 0.42 |

<sup>a</sup>Abbreviations: CSF – cerebrospinal fluid; CNS – central nervous system; ICU – intensive care unit; IQR – interquartile range; WFNS – world federation of neurological surgeons
<sup>b</sup>Statistical tests: Group differences were tested with a Chi-square test for categorical variables and Wilcoxon test for continuous variables.
<sup>c</sup>Secondary open operation defined as craniotomy or craniectomy after completion for treatment of the aneurysm.
<sup>d</sup>CNS infection defined as infection of the central nervous system, meninges, or cerebrospinal fluid (microbiologically proven or clinical signs treated empirically with 4 or more consecutive days of antibiotics).

In the group that received fludrocortisone, the median dosage and the number of patients treated per ICU admission day are plotted in Figure 1. More patients treated with fludrocortisone were also co-treated with enteral salt [13 (32%) vs. 6 (16%); P=0.12] and hypertonic saline [23 (56%) vs. 6 (16%); P<0.01]. When used, the amount of enteral salt and hypertonic saline did not significantly differ between groups (Table 2).

**Table 2.** Sodium supplementation between ICU admission and day 14 in aSAH patients treated with and without fludrocortisone.

| **Sodium Replacement<sup>a</sup>** | **Fludrocortisone (n=41)** | **No Fludrocortisone (n=37)** | **P value** |
| --- | --- | --- | --- |
| **Hypertonic saline** |  |  |  |
| median [IQR], mL | 4500 [1000-6000] | 1625 [1062.50-5187.50] | 0.79 |
| n | 13 (32%) | 6 (16%) | 0.12 |
| **Enteral salt** |  |  |  |
| median [IQR], g | 14 [6-47.5] | 11.5 [7.25-16.5] | 0.40 |
| n | 23 (56%) | 6 (16%) | <0.01 |

<sup>a</sup>Abbreviations: aSAH – aneurysmal subarachnoid hemorrhage; ICU – intensive care unit; IQR – interquartile range

**Figure 1.**
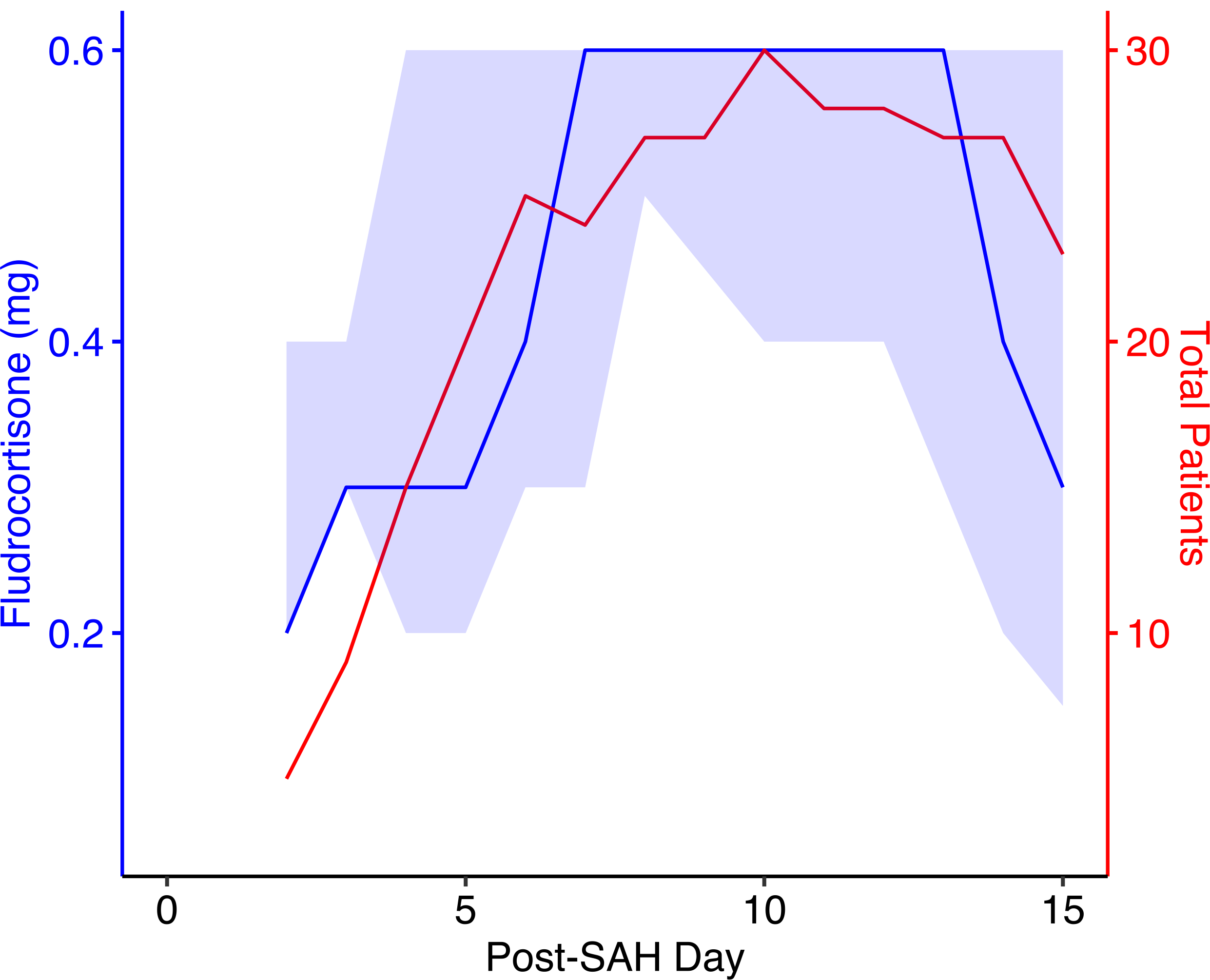
Fludrocortisone use in aneurysmal subarachnoid hemorrhage patients. The median dose of fludrocortisone (blue line) with interquartile range (light blue shade) and the number of patients treated with fludrocortisone (red line) are plotted against the post-subarachnoid hemorrhage day.

### Outcomes

Compared with patients who did not receive fludrocortisone, patients who received fludrocortisone experienced a lower mRS scores at 90 days (adjusted common odds ratio, aOR, of 0.33; 95% CI, 0.14 to 0.80; P=0.02), with values below 1.0 favoring receipt of fludrocortisone (Figure 2; Table 3). Excluding the 6 patients who did not have a mRS score at 90 days resulted in an aOR of 0.36 (95% CI, 0.14 to 0.92; P=0.03). Substituting them with imputed values resulted in an aOR of 0.36 (95% CI, 0.14 to 0.91; P=0.04). Because the use of enteral salt and hypertonic saline correlated with the use of fludrocortisone (Table 3), we introduced these variables in the regression model. Both did not independently correlate with the outcomes (use of enteral salt aOR 0.5 [0.18-1.35]; P=0.17; and use of hypertonic saline aOR 1.30 [0.45-3.70]; P=0.63), but the use of fludrocortisone maintained a similar association as in the primary analysis (aOR 0.41 [0.15-1.04]; P=0.06). Because these patients were enrolled in the SMART trial, comparing the primary use of balanced crystalloids to saline for intravenous fluid therapy, we included the trial fluid assignment as an interaction variable in the model. Of the 41 patients who received fludrocortisone, 21 were assigned saline and 20 were assigned a balanced crystalloid; and of the 37 patients who did not receive fludrocortisone, 20 were assigned saline and 17 were assigned a balanced crystalloid. In this model, the use of fludrocortisone continued to maintain a similar association as in the primary analysis (aOR 0.15 [0.04-0.60]; P=0.01). The use of saline was associated with better outcomes (aOR 0.24 [0.06-0.96]; P=0.05). Importantly, in the model the interaction term of fluid assignment and fludrocortisone did not associate with outcome (aOR 3.22 [0.50-21.6]; P=0.22).

**Table 3.** Ordinal regression models for 90-day modified Rankin Scale (mRS) scores. Adjusted common odds ratios with 95% confidence intervals and corresponding P values are reported.

| Co-variable | Effect size |
| --- | --- |
| Fludrocortisone (vs. no fludrocortisone) | 0.33 [0.14-0.80], P=0.02 |
| Age (continuous) | 1.06 [1.02-1.10], P=0.01 |
| Time to treatment (continuous) | 1.00 [0.91-1.10], P=0.96 |
| Hypertension (no hypertension as reference) | 1.20 [0.44-3.31], P=0.72 |
| Endovascular embolization (clip obliteration as reference) | 1.01 [0.21-4.80], P=0.99 |
| World Federation of Neurological Surgeon's grade (Grade 1 as reference) |  |
| Grade 2 | 2.27 [0.67-7.88], P=0.19 |
| Grade 3 | 4.70 [0.92-24.7], P=0.06 |
| Grade 4 | 5.36 [1.50-20.2], P=0.01 |
| Grade 5 | 18.7 [4.78-79.0], P<0.01 |

**Figure 2.**
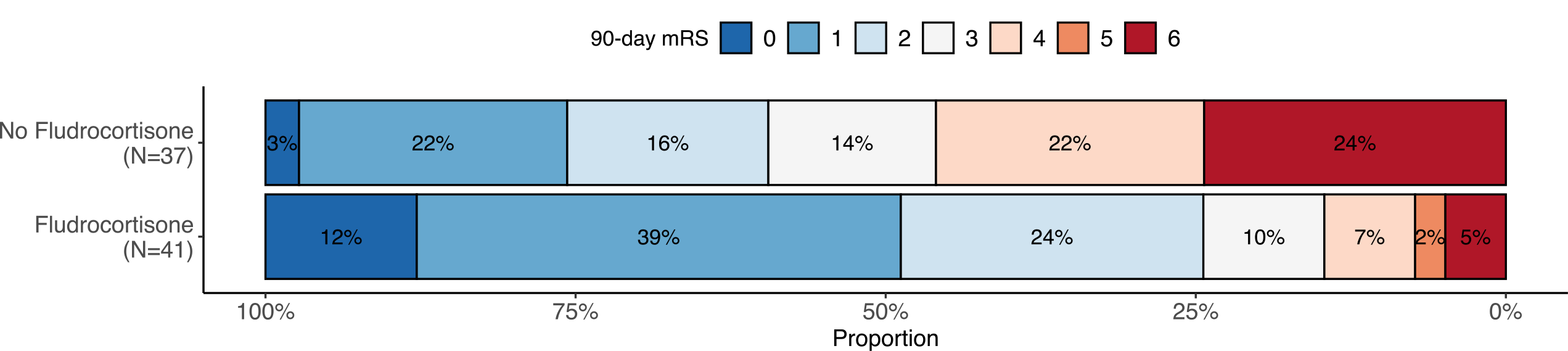
Functional outcomes of patients with aneurysmal subarachnoid hemorrhage treated with and without fludrocortisone. Distribution of scores on the modified Rankin scale (mRS) at 90 days according to the treatment with fludrocortisone. The mRS ranges from a score of 0, indicating no symptoms, to a score of 6, indicating death. The numbers within the bars are percentages of patients who attained the score.

Patients treated with or without fludrocortisone experienced similar incidences of organ dysfunction events (Table 4). Patients treated with fludrocortisone had higher but not statistically significant incidences of symptomatic vasospasm [17 (42%) vs 8 (22%); P=0.10], delayed cerebral ischemia events [17 (42%) vs 8 (22%); P=0.16], and pulmonary edema [6 (15%) vs 3 (8%); P=0.59]. No patient with fludrocortisone had cardiac dysfunction (0 (0%) vs 4 (11%); P=0.10). Length of hospital stay did not differ with the use of fludrocortisone (median 17 days [IQR 14-22] vs 16 [11-20]; Table 4).

**Table 4.** Outcomes.

| Outcome<sup>a</sup> | No Fludrocortisone (n=37) | Fludrocortisone (n=41) | Adjusted common odds ratio (95% CI) | P value<sup>c</sup> |
| --- | --- | --- | --- | --- |
| **Primary outcome** |  |  |  |  |
| mRS at 90 days, *n* | 3 [2-4] | 1 [1-2] | 0.33 [0.14-0.80]<sup>b</sup> | 0.02 |
| **Secondary outcomes** |  |  |  |  |
| *Clinical outcomes, n (%)* |  |  |  |  |
| Length of stay, days | 16 [11-20] | 17 [14-22] |  | 0.39 |
| *Adverse organ-based events, n (%)* |  |  |  |  |
| Symptomatic vasospasm | 8 (22%) | 17 (42%) | - | 0.10 |
| Delayed cerebral ischemia | 8 (22%) | 16 (39%) | - | 0.16 |
| Pulmonary edema<sup>d</sup> | 3 (8%) | 6 (15%) | - | 0.59 |
| Cardiac dysfunction<sup>e</sup> | 4 (11%) | 0 (0%) | - | 0.10 |

<sup>a</sup>Abbreviations: CI – confidence interval; mRS – modified Rankin Scale
<sup>b</sup>Adjusted common odds ratio from a proportional odds regression analysis
<sup>c</sup>Group differences were tested with a Chi-square test for categorical variables and Wilcoxon test for continuous variables; if odds ratios are presented, associated P values are presented
<sup>d</sup>Pulmonary edema evidenced by a chest radiograph and/or initiation of a diuretic treatment with the goal of treating pulmonary edema.
<sup>e</sup>Cardiac dysfunction defined as myocardial infarction, new onset of heart failure, or initiation of inotropic therapy.

## DISCUSSION

The 2023 American Heart Association aSAH guidelines recommend that “the use of mineralocorticoids is reasonable to treat natriuresis and hyponatremia,”(12) citing its efficacy from randomized controlled trials (8). Natriuresis-driving hypovolemia can compromise organ perfusion, risking life-threatening ischemic strokes (in the setting of cerebral vasospasm), AKI, and cardiopulmonary dysfunction (11). Indeed, a meta-analysis showed that fludrocortisone reduces symptomatic vasospasm in aSAH patients (8). A 8346-patient population-based study showed that poor hemodynamic status (hyponatremia and hypovolemia) increases the risk of adverse events from vasospasm (17) resulting in poor outcomes. A similar therapeutic effect of fludrocortisone has been observed in patients with TB meningitis who also exhibit natriuresis-driven hypovolemia and strokes (18). However, its impact on outcomes of aSAH patients is unclear and has not been reported since 1999 (19). In this retrospective study, we found that fludrocortisone therapy was independently associated with better outcomes following aSAH.

The results of our study must be interpreted within its limitations. First, this study has inherent retrospective bias. Second, the effect size we observed is large and may be due to the study’s small sample size. The sample size was selected based on the identification and selection of a *consecutive* cohort of aSAH patients, which was facilitated by the SMART trial. Third, conduct at a single center limits generalizability. Fourth, fludrocortisone was not protocolized and was often started by some providers after the patient exhibited symptomatic vasospasm and hypovolemia, weakening any causal link between the outcomes and fludrocortisone. Thus, the higher rate of symptomatic vasospasm and delayed cerebral ischemia seen in the group treated with fludrocortisone should be interpreted with great caution. All prior studies, including randomized controlled trials, have shown fludrocortisone is associated with lower incidences of symptomatic vasospasm and delayed cerebral ischemia (8). However, it is noteworthy that despite greater incidences of symptomatic vasospasm and delayed cerebral ischemia, the use of fludrocortisone in these patients was associated with better outcomes. Our results must be confirmed with prospective randomized controlled trials.

## CONCLUSIONS

In this retrospective cohort analysis, the use of fludrocortisone was associated with better outcomes in aSAH patients. This finding must be confirmed with a randomized controlled trial.

## Data Availability

All data produced in the present study are available upon reasonable request to the authors

